# Menstruation at high altitude: results from an opportunistic cross-sectional survey of Andean women living above 3500 m

**DOI:** 10.1101/2025.08.15.25333517

**Authors:** Alejandro Correa-Paris, Veronica Gorraiz Ochoa, Alfonso Correa Uribe

## Abstract

**Introduction:** The menstrual cycle characteristics are essential to sexual and reproductive healthcare. Exposure to high altitudes reduces blood oxygen pressure and may result in hypoxia and mountain sickness. Data concerning its impact on menstruation is lacking.

**Methods:** We conducted an opportunistic cross-sectional survey to study the menstrual cycles and the prevalence of dysmenorrhea in residents living above 3500 m in the Andean mountains. We measured dysmenorrhea on a 10-grade scale and evaluated bleeding based on the validated menstrual bleeding questionnaire. The covariates included socio-demographical, anthropometrical, and medical data.

**Results:** one hundred thirty-six subjects from Argentina (6%), Bolivia (39%), and Peru (55%) were included (81% response rate). Most women had regular menstrual cycles and menarche at 13 years of age. Seven out of ten participants did not use contraception. The median menstrual pain score was 3.9 (interquartile range 2.8) with an estimated population mean of 3.7 ± 0.4, but 23.6% of participants had moderate to severe dysmenorrhea (grade 2-3). Dysmenorrhea and heavy menstrual bleeding were more prevalent with increasing altitude. There was no significant association between parity or breastfeeding and dysmenorrhea, while the use of hormonal contraception significantly decreased the severity of pain. Nearly 20% considered their menstruation normal despite having symptoms consistent with heavy menstrual bleeding. All women denied having any known gynecological illnesses. Heavy menstrual bleeding and severe dysmenorrhea were inversely associated with the education level. Only 22.8% had previously sought medical attention because of dysmenorrhea, 42% needed to take analgesics, and most preferred natural remedies such as herbal preparations (62%).

**Discussion:** Our findings reflect a lack of menstrual education and underdiagnosis of gynecological—especially uterine— disorders in these rural and remote populations. Various coping mechanisms (endogenous antioxidant activity, dietary antioxidants, genetic modifications, etc.) can explain the adaptation and low intensity of pain, but the impact of high altitude and hypoxia on sexual and reproductive health is complex and poorly understood.

**Conclusions:** Andean high-altitude residents (living above 3500 m) have delayed menarche, regular menstrual cycles, and low average menstrual pain. However, the prevalence of moderate to severe dysmenorrhea and heavy menstrual bleeding is high and should be studied in more depth. High altitude appears to influence menstrual pain and bleeding; this should be evaluated further in a larger controlled trial comparing sea-level with high Andean residents. More studies are needed to understand the needs of this special population in terms of menstrual education and sexual and reproductive healthcare.

## INTRODUCTION

The characteristics of the menstrual cycle reflect normal reproductive function and are an essential guide for sexual and reproductive healthcare. The presence of pathological symptoms (e.g., intense pain, heavy menstrual bleeding) can compromise quality of life and suggest an underlying disease with potentially serious health consequences.

Dysmenorrhea is the most frequent symptom affecting women’s health worldwide[1]. Whether primary or secondary, it is a public health issue: it is the leading cause of absenteeism (work or school) in young women[2,3].

Humans have settled in nearly all habitable locations on the planet. Exposure to high altitudes reduces blood oxygen pressure and may result in hypoxia. This can lead to acute (AMS) or chronic (CMS) mountain sickness, which can lead to various complications, including cardiopulmonary and obstetric disorders[4], but humans have successfully adapted to live at high altitudes in all continents except Europe and Oceania.

Mountain sickness (MS) usually appears above 2500 m, but CMS is more common between 3500 and 4000 m[5,6]. The prevalence of CMS is higher in the Andes, rare in the Himalayas, and practically nonexistent in Ethiopia[7]. Overall, CMS affects more women than men[5].

There has been some interest in investigating the effects of high-altitude hypoxia on reproductive physiology, but most studies have focused only on reproductive and obstetric outcomes[8–11]. Very few studies have analyzed variations in menstrual physiology in the hypoxic environment of the highlands.

The only available data regarding dysmenorrhea at high altitudes are from a study conducted in Peru in 1993 that compared residents at sea level (Lima) with those at high altitudes (Cerro de Pasco). At nearly 4300 m, the most frequently reported cause for gynecological consultation was dysmenorrhea (45%), but at sea level, it was the least frequent motive (10%)[8].

There are limited data available on the characteristics and variations of the menstrual cycle in high-altitude residents. There are no published data concerning the characteristics of menstrual bleeding, the severity of dysmenorrhea, or the consequences of and attitudes towards these symptoms.

We conducted a cross-sectional survey to describe the characteristics of the menstrual cycle and the prevalence of dysmenorrhea in residents living above 3500 m above mean sea level (AMSL) in the Andean mountains.

This is the first study to describe the menstrual cycle characteristics of high-altitude residents across the Andean plateau.

## METHODS

We conducted an opportunistic cross-sectional survey between August 2019 and March 2020 in all available and permanently inhabited settlements above 3500 m above mean sea level (AMSL) in the South American Andes mountain range.

### Materials

Surveys were conducted by a single person (ACP, VGO, or BC), mainly using written surveys on-site (Supplementary material S2: survey). Participants were also offered the option to complete the survey online using their mobile phones via a QR code.

The survey was designed to include four groups of questions, with information related to demographics, medical history and lifestyle, previous gynecological and obstetrical history, and menstrual cycle characteristics. The survey was internally and externally validated after peer review by an experienced gynecologist and researcher (ACU), an external epidemiologist, and a Bolivian physician (BC, for comprehension and proofreading).

Women were invited to participate in an anonymous survey to study women’s health in high altitudes. Those who accepted were given an information sheet with a description of the study and instructions to complete the survey.

Participants completed a two-page anonymous questionnaire without receiving any help or additional indications. If someone was unable to fill out the written survey, the surveyor read the questions aloud and filled in the answers as given literally by the participant.

### Participants

We included all women of reproductive age who were permanent residents at an altitude greater than or equal to 3500 m AMSL. Permanent residents were defined as having lived in that location for at least 6 months (since adaptation to high altitude usually occurs after that time).

The inclusion criteria were: age between 16 and 45 years, and having a menstrual period. The exclusion criteria were: amenorrhea (>3 months), current pregnancy, menopause, and inability to complete the survey.

### Study protocol

The study protocol planned to visit several rural and urban locations in the Andes in Argentina, Chile, Bolivia, Peru, Ecuador, and Colombia, covering the main localities above 3500 m AMSL.

The route was mapped according to accessibility by motor vehicle. Still, it was not scheduled for any particular day, as unexpected incidents are frequent on Andean roads (roadblocks, protests, extreme climate, etc.).

We visited all permanent human settlements accessible by car and surveyed all available female residents (in public spaces, on the streets, or in door-to-door encounters). The sampling method was opportunistic.

### Measurements

Altitude was measured on-site using a GPS in meters (m) above mean sea level (AMSL). In case the location had variable altitude (i.e., several areas or neighborhoods at different elevations), we used the official mean altitude as stated by the government.

The main outcome measure was the dysmenorrhea intensity score. We used the validated 10-grade graphic rating scale (GRS) to determine the dysmenorrhea intensity score (0 to 10), which is easier to understand than the traditional visual analog scale (VAS) and yields fewer erroneous interpretations and false 0 values[12]. We also used a verbal multidimensional system to classify the severity of dysmenorrhea (grades 0 to 3, see Figure 4) based on the patient’s symptoms[13].

The secondary outcomes included: other characteristics of dysmenorrhea (i.e., need for analgesia), menstrual bleeding and cycle characteristics, lifestyle and anthropometric characteristics, including the frequency of physical activity and main mode of transportation, past medical and gynecological history, and demographic characteristics, including age and education. All studied variables are listed in (Supplementary material: Table S1).

We included questions related to menstrual bleeding volume to evaluate the presence of abnormal or heavy menstrual bleeding (HMB), according to the definitions of the International Federation of Gynecology and Obstetrics (FIGO) [14]. We defined 4 sentences based on the validated menstrual bleeding questionnaire (MBQ)[15], a patient-reported outcome instrument for heavy menstrual bleeding. We included only 4 questions related to HMB (instead of all 16 in the MBQ) to keep the questionnaire short and easy to complete, and because the main outcome was not heavy menstrual bleeding (Supplementary material S2: survey). We defined a composite outcome measure as an objective marker of heavy menstrual bleeding defined by the presence of any of the 3 HMB statements considered abnormal and/or having a menstrual period of more than 7 days.

### Analyses

Incomplete surveys (if more than 80% of questions were left unanswered) were excluded from the analysis.

The sample size was calculated in advance using the Granmo calculator software (v7.12) and based on the results of a study describing the prevalence and severity of dysmenorrhea[16] where the mean score was 6.79 ± 2.64 on a 10-grade VAS. On the basis of these results, we determined that a random sample of 119 individuals would be sufficient to estimate the population mean with a precision of ± 0.5, standard deviation of 2.64, and 95% confidence. The anticipated replacement rate was estimated to be 10%.

For the descriptive analysis, we calculated percentages, medians, interquartile range (IQR), and standard error of the mean (SEM) when appropriate. We used statistical tests as per the distribution of data (normal vs. non-normally distributed), the Chi-square or Fisher tests for qualitative variables, and the Mann-Whitney U or t-tests based on the nature of the quantitative variables.

Statistical analysis was performed using R software version 4.0 (R Core Team, GNU) and Wizard Statistics & Analysis version 1.9. Statistical significance was defined as p < 0.05.

### Ethics approval

The study protocol was approved by the Institutional Review Board (IRB) of HIRU (Project Evaluation Committee) with protocol n° PI-2019/08-01. Although the study was planned to be conducted in different countries, additional IRB approval was waived after consulting with a member of an external Ethics Committee in Colombia. This was because the study was descriptive, compliant with the Declaration of Helsinki, and based on a voluntary and anonymous questionnaire. The study was reported in accordance with the STROBE (Strengthening the Reporting of Observational Studies in Epidemiology) guidelines (Supplementary material S3: STROBE checklist).

## RESULTS

We were only able to sample residents in Argentina, Bolivia, and Peru. When we visited locations in Chile, women declined to participate. The data collection period of our study was interrupted by the COVID-19 pandemic; thus, it was not possible to collect information from Ecuador and Colombia.

The response rate was 81% (136/168); most answers were collected from written surveys (95%), and we did not exclude any surveys because of missing data (Figure 1).

**Figure 1.**
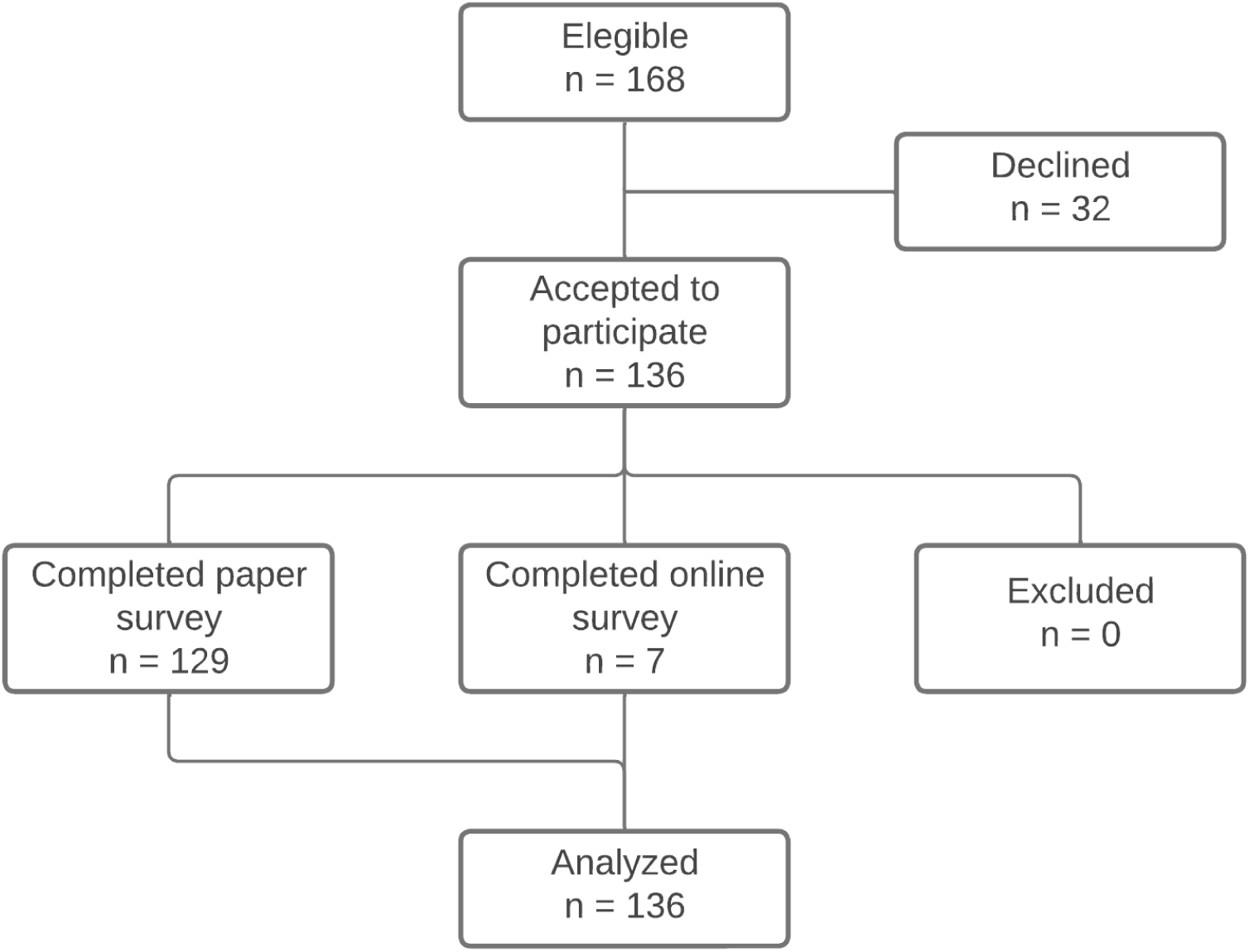
Study flowchart

### Main sample characteristics

We analyzed a total of 136 participants. Most of them were young and healthy individuals with no relevant medical history and normal BMI (Tables 1 and 2).

**Table 1.**
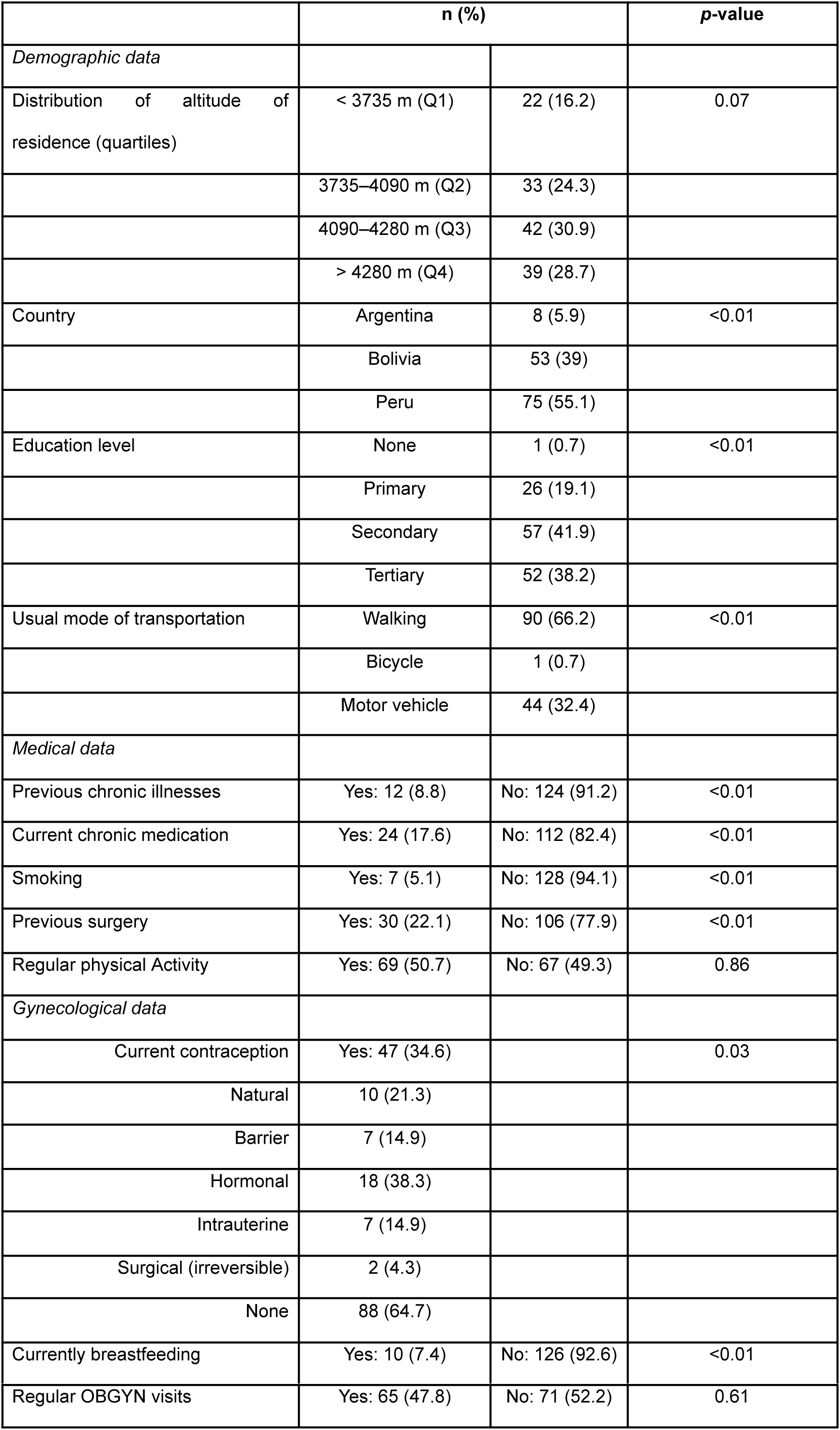
Descriptive analysis and characteristics of our sample

|  | n (%) |  | p-value |
| --- | --- | --- | --- |
| *Demographic data* |  |  |  |
| Distribution of altitude of residence (quartiles) | < 3735 m (Q1) | 22 (16.2) | 0.07 |
|  | 3735–4090 m (Q2) | 33 (24.3) |  |
|  | 4090–4280 m (Q3) | 42 (30.9) |  |
|  | > 4280 m (Q4) | 39 (28.7) |  |
| Country | Argentina | 8 (5.9) | <0.01 |
|  | Bolivia | 53 (39) |  |
|  | Peru | 75 (55.1) |  |
| Education level | None | 1 (0.7) | <0.01 |
|  | Primary | 26 (19.1) |  |
|  | Secondary | 57 (41.9) |  |
|  | Tertiary | 52 (38.2) |  |
| Usual mode of transportation | Walking | 90 (66.2) | <0.01 |
|  | Bicycle | 1 (0.7) |  |
|  | Motor vehicle | 44 (32.4) |  |
| *Medical data* |  |  |  |
| Previous chronic illnesses | Yes: 12 (8.8) | No: 124 (91.2) | <0.01 |
| Current chronic medication | Yes: 24 (17.6) | No: 112 (82.4) | <0.01 |
| Smoking | Yes: 7 (5.1) | No: 128 (94.1) | <0.01 |
| Previous surgery | Yes: 30 (22.1) | No: 106 (77.9) | <0.01 |
| Regular physical Activity | Yes: 69 (50.7) | No: 67 (49.3) | 0.86 |
| *Gynecological data* |  |  |  |
| Current contraception | Yes: 47 (34.6) |  | 0.03 |
| Natural | 10 (21.3) |  |  |
| Barrier | 7 (14.9) |  |  |
| Hormonal | 18 (38.3) |  |  |
| Intrauterine | 7 (14.9) |  |  |
| Surgical (irreversible) | 2 (4.3) |  |  |
| None | 88 (64.7) |  |  |
| Currently breastfeeding | Yes: 10 (7.4) | No: 126 (92.6) | <0.01 |
| Regular OBGYN visits | Yes: 65 (47.8) | No: 71 (52.2) | 0.61 |

**Table 2.** Summary of descriptive statistics.

|  | Median (range) | IQR | SEM |
| --- | --- | --- | --- |
| *Demographic data* |  |  |  |
| Age (years) | 28 (15–45) | 10 | 0.64 |
| Residence altitude (m) | 4090 (3640–4700) | 545 | 26.7 |
| Weekly exercise frequency | 2 (0.3–7) | 2 | 0.2 |
| BMI (kg/m<sup>2</sup>) | 24.2 (16.4–40.2) | 4.6 | 0.34 |
| *Medical data* |  |  |  |
| Menarche (years) | 13 (11–18) | 2 | 0.13 |
| Pregnancies (n) | 1 (0-9) | 2 | 0.15 |
| Abortions (n) | 0 (0–6) | 0 | 0.06 |
| Parity (n° of children) | 1 (0-5) | 2 | 0.13 |
| Dysmenorrhea score (GRS) | 3.9 (0–10) | 2.8 | 0.2 |

IQR: interquartile range, SEM: standard error of the mean, GRS: Graphic rating scale (0–10)

The sample included significantly more women from Peru (55%), and Argentina accounted for the smallest proportion (5.9%) (Table 1, Supplementary figure S4).

The median altitude was evenly distributed but varied according to the country. Residential altitude was unequally distributed (different quartile proportions, p = 0,013). Based on the country of residence, all Argentinians lived between the 2nd and 3rd quartiles of altitude (e.g., 3735–4280 m AMSL). Only women from Peru lived above 4280 m (52% of them) and were in all altitude quartiles. The country of residence did not influence the following variables: age, physical activity, main mode of transportation, or most medical history variables. The vast majority of the sample was non-smokers (6 of the 7 women who smoked lived in Bolivia). Two-thirds of our sample chose walking as their customary mode of transportation (66.7%); this proportion varied according to country and was lowest in Bolivia (45.3%).

Education levels varied according to the country of residence. Most Peruvian and Argentinian women had a secondary education (46.7% and 62.5% respectively), whereas most Bolivian women in our sample had a superior education (60.4%).

### Obstetric and gynecological history

The age at menarche (median 13 years, min 11, max 18, IQR 2) was not influenced by altitude in our sample, but it varied according to the country of residence. Argentinians were the oldest (median 14.5 years), followed by Peruvians (14 years) and Bolivians (13 years). Most participants had few children, and the majority had never had any abortions (85%) (Table 2).

There was no correlation between the main outcome measure and past pregnancies or parity (comparison by quartiles: Kruskal-Wallis p=0.84 and p=0.64, respectively). We found similar results when comparing dysmenorrhea grade with gestations and parity (chi-squared p=0.68 and p=0.47, respectively).

More than half of the participants did not use any contraception, and among users, the most common method was hormonal (38%), followed by natural methods (21%) (Table 1). Less than half of the participants regularly visited obstetrics and gynecology specialists (47.8%).

### Menstrual symptoms

Most participants had regular menstrual cycles and normal uterine bleeding (Table 3).

**Table 3.** Characteristics of the menstrual cycle and symptoms

|  |  | n | % | *p*-value |
| --- | --- | --- | --- | --- |
| Cycle duration (days) | <21 | 25 | 18.4 | <0.01 |
|  | 21–35 | 104 | 76.5 |  |
|  | >35 | 7 | 5.1 |  |
| Bleeding days | ≤7 | 129 | 94.9 | <0.01 |
|  | 8-14 | 7 | 5.1 |  |
|  | >14 | 0 | 0 |  |
| *Bleeding quantity* | Subjectively normal | 99 | 72.8 | <0.01 |
|  | Subjectively abnormal | 37 | 27.2 |  |
| Number of HMB indicators <sup>†</sup> | 1 | 35 | 25.7 | <0.01 |
|  | 2 | 15 | 11 |  |
|  | 3 | 4 | 2.9 |  |
| *Menstrual pain severity* | None | 43 | 31.6 | <0.01 |
|  | Grade 1 | 60 | 44.1 |  |
|  | Grade 2 | 22 | 16.2 |  |
|  | Grade 3 | 10 | 7.4 |  |
| Previously sought medical aid for pain | Yes | 31 | 22.8 | <0.01 |
|  | No | 105 | 77.2 |  |
| Analgesia | Yes | 57 | 41.9 | <0.01 |
|  | Self-medication | 44 | 75.9 |  |
|  | Prescribed | 14 | 24.1 |  |
|  | No | 79 | 58.1 |  |
| Type of analgesia | NSAIDs alone | 13 | 24.5 | <0.01 |
|  | NSAID + other | 2 | 3.8 |  |
|  | non-NSAID analgesics | 4 | 7.5 |  |
|  | Herbal | 33 | 62.3 |  |
|  | Other | 1 | 1.9 |  |

HMB: Heavy menstrual bleeding, NSAID: Non-steroidal anti-inflammatory drug.
† The survey included three statements indicating possible HMB: 1) “I need to change the menstrual protection (pad, tampon, etc.) every hour or more frequently”, 2) “I need to use more than a simple protection or more than one type of protection” (e.g. “I need to use a double pad”, or “I need to use a special diaper”), 3) “I need to wake up at night to change my menstrual protection because of the bleeding”; subjects could mark all that applied.

The main outcome measure was independent of all secondary variables except contraception, use of analgesia for dysmenorrhea, previous medical consultation for dysmenorrhea, and variables defining heavy menstrual bleeding (HMB).

We evaluated the study variables by comparing the use of hormonal contraceptives vs. no contraception or non-hormonal methods. The proportion of women without dysmenorrhea (grade 0) was higher among those who used hormonal contraception (61.1% vs. 27.4%, p=0.04). Similarly, the median dysmenorrhea score was lower in those using hormonal contraception (2.1 vs 4.0, p=0.02).

More than two-thirds of women (68.4%) did not report any pain during menses. Nearly one out of four (23.7%) participants reported severe dysmenorrhea (grade 2 or 3). The median dysmenorrhea score was 3.9 (IQR 2.8), with an estimated population mean of 3.7 ± 0.4.

We found a low positive correlation between altitude and the intensity of menstrual pain (Spearman rho = 0.21, p = 0.01). When analyzing the dysmenorrhea score at different altitude groups (quintiles), we observed a trend of increasing pain at higher altitudes (Figure 2).

**Figure 2.**
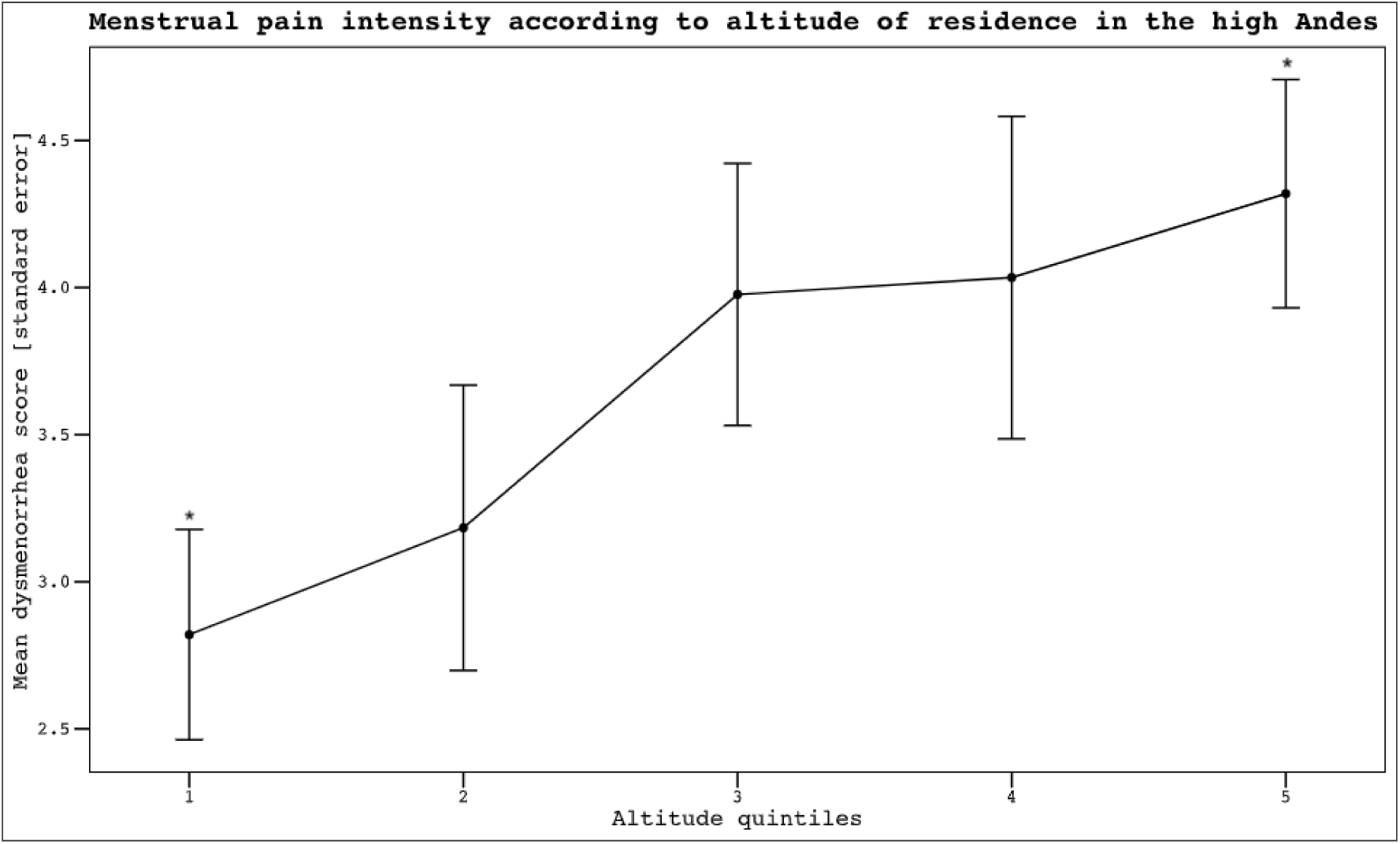
Menstrual pain intensity according to the altitude of residence. The mean pain score for menstrual pain is based on the 10-grade graphic rating scale (GRS), and the brackets represent the standard error. * *p* < 0.05

There was no significant association between parity and dysmenorrhea. Women who were breastfeeding at the time of the survey (n=10) did not show any significant differences in dysmenorrhea (GRS 2.9 vs 4.0, p=0.42) or menstrual blood loss (no differences in questions regarding heavy menstrual bleeding, p>0.4).

Subjects who stated that their menstrual bleeding was not normal showed higher dysmenorrhea scores than women who considered their bleeding to be normal (4.2 vs. 3.2, p<0.001). Women who marked the statements related to heavy menstrual blood loss showed increasing dysmenorrhea scores corresponding to the number of positive questions (Figure 3). The median dysmenorrhea score was 2.9 if no HMB statements were marked, and with an increasing number of HMB questions answered (1, 2, or 3), the medians were 4.1, 4.0, and 6.1, respectively (p=0.02).

**Figure 3.**
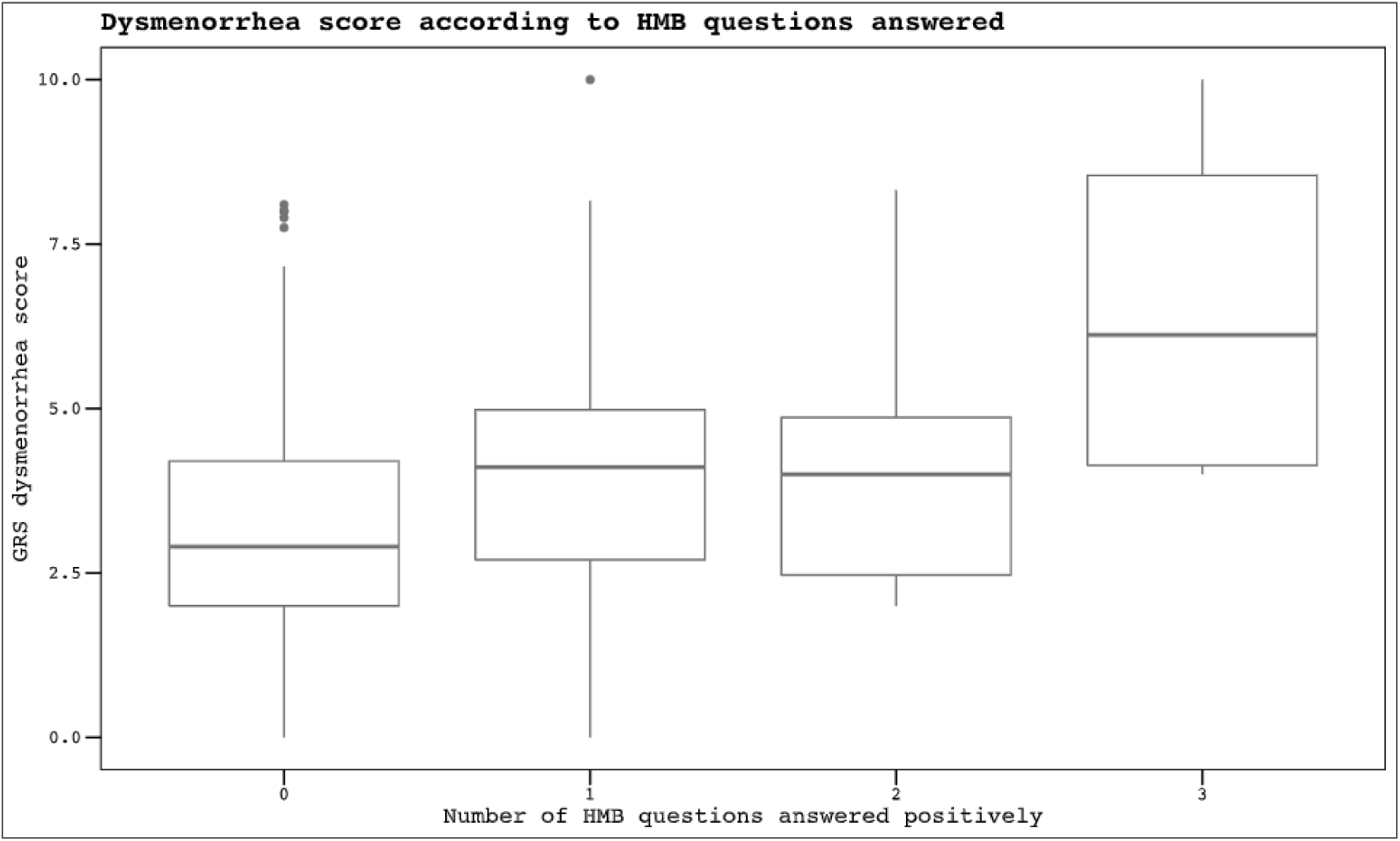
Boxplot of dysmenorrhea score according to answers to heavy menstrual bleeding questions. HMB: heavy menstrual bleeding, GRS: 10-grade graphic rating scale. The boxes show the mean and 95% confidence interval GRS score of menstrual pain according to the number of affirmatively answered questions consistent with HMB. In the survey, participants were asked to mark all that applied. The three HMB statements were: I need to change protection (towel, pad, or tampon) every hour or less; I need to use more than one protection (double towel, diaper, etc.); I need to get up at night to change protection because of bleeding. The dots represent outliers. *p* = 0.03

Only 22.8% of women had previously sought medical attention because of dysmenorrhea. This group of women showed significantly higher dysmenorrhea scores (4.2 vs. 3.2, p=0.017).

Although the overall median dysmenorrhea score was low (3.9), nearly 42% of the women needed to take analgesics.

Most women who used analgesia opted for natural remedies such as herbal preparations (62%), and less than 25% of analgesics were prescribed by a health care professional (Table 3). We observed that the severity of menstrual pain decreased with increasing education (Figure 4).

**Figure 4.**
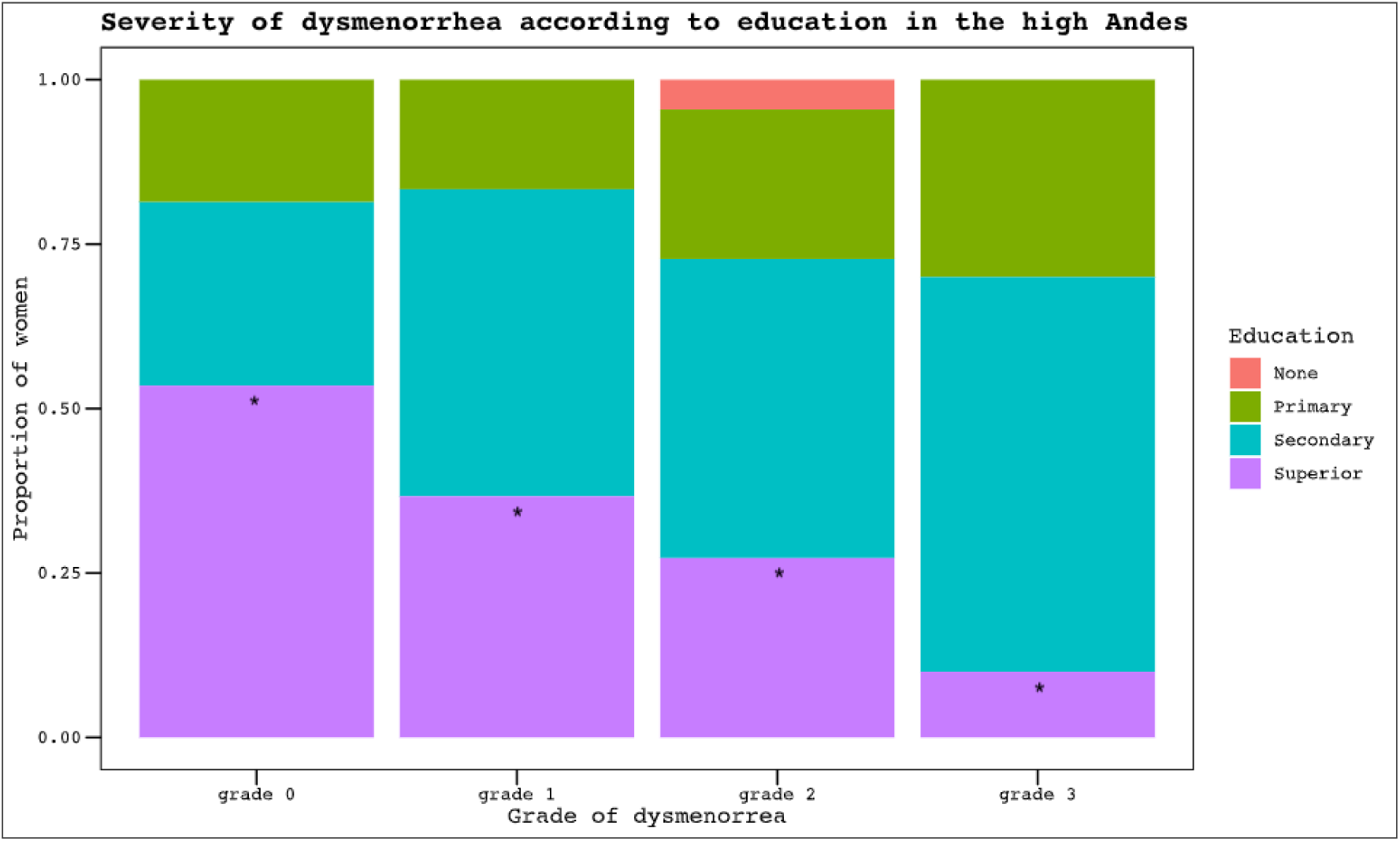
The severity of dysmenorrhea according to education level in the high Andes. The grade of dysmenorrhea is a multidimensional system to classify the severity of menstrual pain described by Andersch and Milsom[13]. Grade 0: Menstruation is not painful, and daily activity is unaffected. Grade 1: Menstruation is painful but seldom inhibits the woman’s normal activity. Analgesics are seldom required. Mild pain. Grade 2: Daily activity is affected. Analgesics are required and effective to avoid absenteeism (work or school). Moderate pain. Grade 3: Activity is inhibited. Poor effect of analgesics. Vegetative and systemic symptoms are present (e.g., headache, tiredness, nausea, vomiting, and diarrhea). Severe pain. * *p* = 0.03

### Heavy menstrual bleeding (HMB)

According to the answers, between 59% and 84% of the subjects experienced some degree of HMB symptoms. The HMB composite outcome was more frequent with increasing altitude (Figure 5) (p=0.02). We also found that the higher the education, the lower the likelihood of having HMB (p=0.03) (Figure 6).

**Figure 5.**
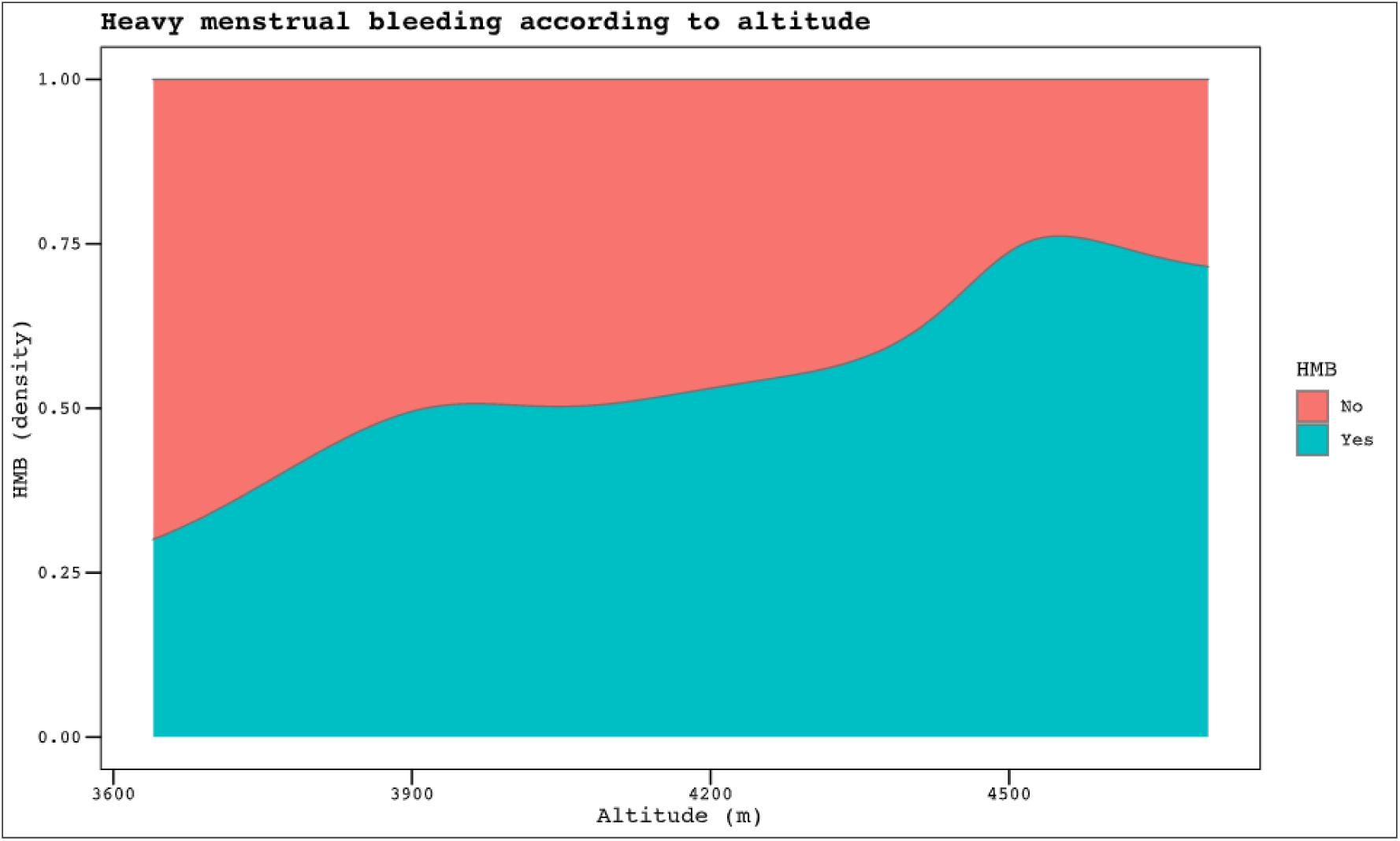
Heavy menstrual bleeding according to altitude. HMB: heavy menstrual bleeding *p* = 0.04

**Figure 6.**
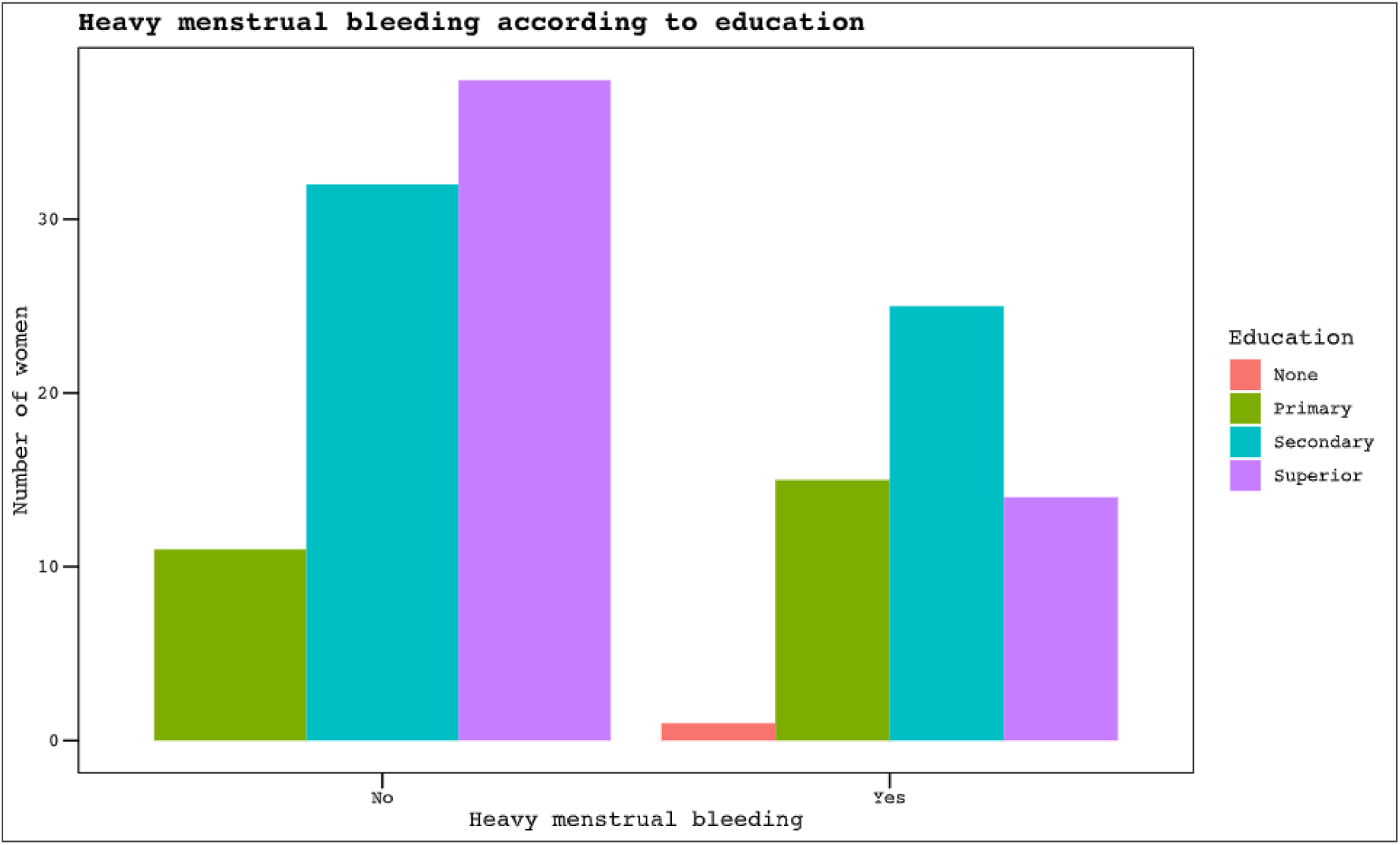
Heavy menstrual bleeding according to education. *p* = 0.03

The anthropometric (e.g., BMI) and lifestyle variables (physical activity and mode of transportation) did not have a significant association with HMB. In addition, past gynecological and obstetrical history did not affect HMB.

The presence of HMB was significantly associated with an increasing grade of dysmenorrhea (p<0.001). Women with HMB had a significantly higher dysmenorrhea score (4.2 vs. 2.8, p=0.05) (Figure 7).

**Figure 7.**
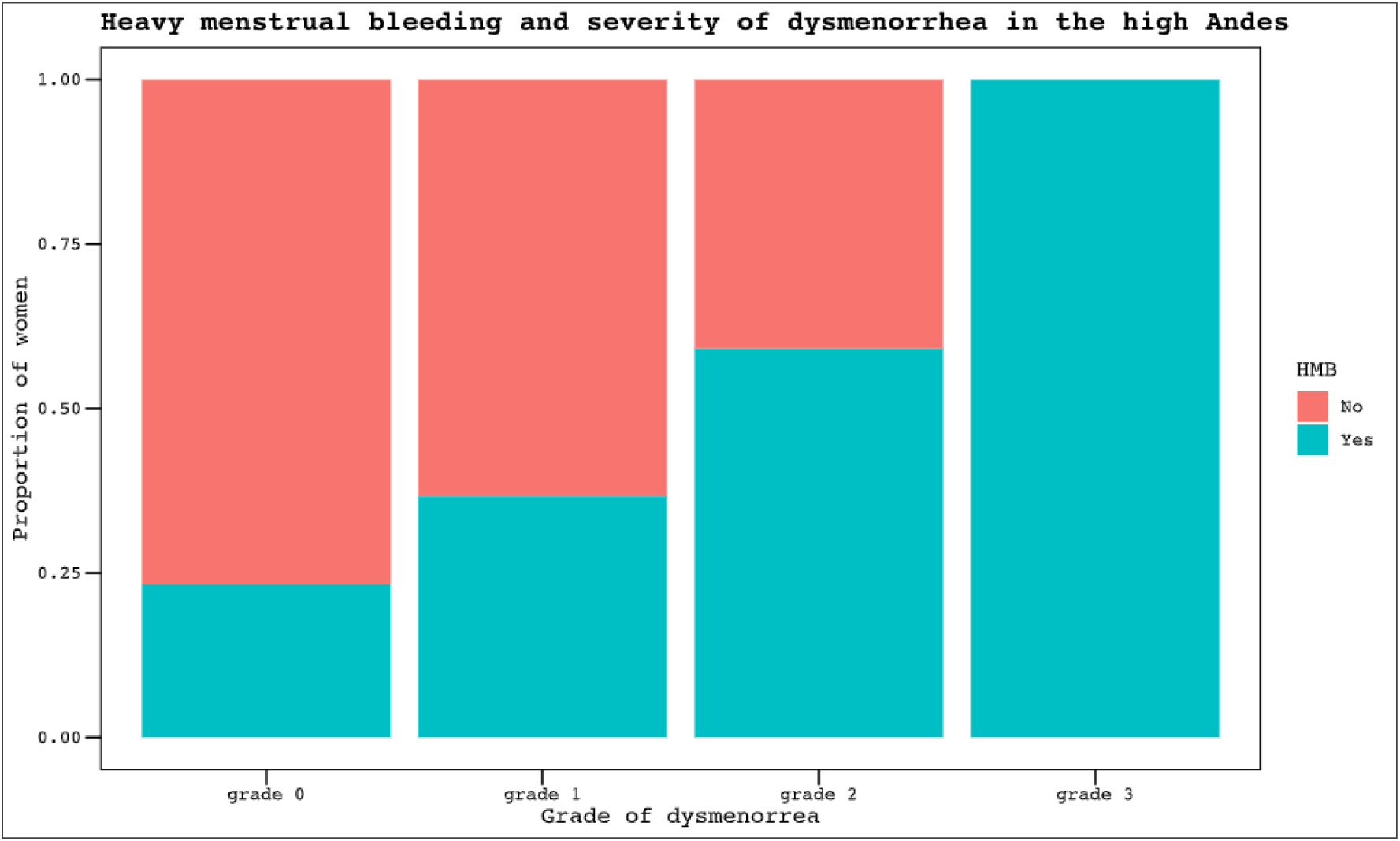
Heavy menstrual bleeding and severity of dysmenorrhea in the high Andes. HMB: heavy menstrual bleeding The dysmenorrhea grading system described by Andersch and Milsom[13] is detailed in Figure 4. *p* <0.01

Nearly 1 out of 5 women (18.2%) claimed their menstrual period was normal, although they had objective signs of heavy menstrual bleeding (having a period for more than 7 days and/or answering affirmatively to 1-3 questions corresponding to HMB according to the MBQ) (chi-squared, p=0.02).

## DISCUSSION

We found that women of reproductive age residing at high altitudes in the Andes have mostly regular menstrual cycles. The average menstrual pain score was medium-low (median 3.9, IQR 2.8), but 23.6% of participants had moderate to severe dysmenorrhea (Table 3). Nearly 7 out of 10 surveyed women did not use contraception, and the most frequent type among users was hormonal methods, followed by natural methods. Online participation was poor (5.15% of responses), probably because of limited internet access in these mostly rural areas.

There was no significant association between parity and dysmenorrhea, despite robust evidence of an inverse relationship between the number of live births and the risk of dysmenorrhea [1]. Notably, nulliparas accounted for only 41.9% of participants.

Nearly 20% of the participants considered their menstruation normal despite having symptoms consistent with heavy menstrual bleeding. This could reflect a lack of menstrual education regarding normal symptoms and duration of menses; in fact, we found that the level of education was inversely proportional to the presence of HMB. It could also reflect the social normalization of pathological menstrual symptoms, possibly due to a lack of access to sexual and reproductive healthcare services or other ethnocultural beliefs[17].

Almost one out of three participants (27%) complained of having abnormal menstrual blood loss, and they also showed significantly more menstrual pain (compared with women who considered their blood loss normal). Similarly, we found increasing dysmenorrhea scores with every affirmative question related to HMB (Figure 7).

Remarkably—and despite our findings regarding severe dysmenorrhea and HMB— none of the participants reported having any gynecological illnesses. However, the prevalence of benign pathologies, such as uterine leiomyomas/fibroids, endometrial polyps, or endometriosis, is known to affect at least 10% of women and can be as high as 70-80%[18–20]. This could indicate undiagnosed uterine pathology, given their high prevalence.

We presumed that there would be a higher prevalence of dysmenorrhea based on the only available data from high-altitude Andean residents. In their 1993 study, Gonzales et al.[8] found that dysmenorrhea was the most frequent motive for consultation with the gynecology service at the local hospital in Cerro de Pasco, Peru (located at 4340 m AMSL) (45.4% vs 10.3% at sea level in Lima). In our cohort (median altitude 4090 m AMSL), 68% of participants claimed they had some menstrual pain, but only 22.8% had ever sought medical aid for this reason. The heterogeneity of our sample (subjects from different countries and ethnic backgrounds) may explain this discrepancy.

The Andes is an extensive mountain range, and although it has been inhabited for thousands of years by different populations, the cultural evolution of each South American country has impacted the genetic diversity of its heritage through migrations and the mixing of people from different origins (e.g., diasporas from Europe and Asia). This can surely influence the adaptation of different people to high altitudes. Chronic exposure to low oxygen levels at high altitudes harms health (Monge’s disease or CMS)[11,21,22], but the consequences on the menstrual and hormonal cycles are not yet well understood. Our results do not support a higher prevalence of menstrual alterations potentially related to high-altitude hypoxia. However, this needs to be specifically addressed in a comparative study (e.g., population at sea level vs. high altitude).

Women residing at high altitudes have been shown to experience menarche at a later age than those at lower altitudes, a phenomenon attributed to altitude-related factors, independent of nutritional status or socio-cultural background[8]. In our sample, the median age of onset of menses was 13 years, which is similar to published data. Similarly, menopause occurs at an earlier age in high-altitude populations, resulting in a shorter reproductive period[23]. A study of women residing above 4300 m in Peru found that lower estrogen levels were associated with lower pulse oxygen saturation[24], which could explain the earlier onset of menopause. An observational study published in 2023 found that high-altitude residents from Peru (at Cerro de Pasco) had significantly worse menopausal symptoms than sea-level residents, hence supporting the hypoxic theory affecting reproductive health[25]. However, other studies have failed to demonstrate any significant differences in the menstrual cycle of residents at sea level and high altitudes[9,26]. Thus, the effect of lower blood oxygen levels on menstrual symptoms remains unclear.

The impact of high altitude on fertility is complex. Historical records had suggested lower fertility presumably due to hypoxia, but more recent studies found conflicting results[27–29]. Previous studies have found lower levels of estrogen, progesterone, and prolactin at sea level, as well as higher FSH levels at high altitudes [8,9,24]. Yet there are no apparent significant differences in the menstrual cycle with increasing altitude. This may reflect the coping capacity of people exposed to chronic hypoxia at high altitudes. Genetic modifications and sociocultural differences have also been demonstrated to influence adaptation to high altitude[11,23].

Some studies have found physical activity to be beneficial for dysmenorrhea[30,31], but in our study, regular exercise and daily walking (as the main mode of transportation, for 66.7% of our sample) did not impact the outcomes.

Other authors have argued that oxidative stress, worsened by altitude, may harm reproductive health[32]. Julian et al. studied oxidative stress in high-altitude pregnant Andean women and highlighted the importance of gene-environment interaction. They demonstrated that endogenous antioxidant activity was greater in Andean women during pregnancy at high but not low altitudes (compared with European controls), and this prevented the appearance of small for gestational age (SGA) newborns, which is more common in the highlands[33]. But it is not clear whether this antioxidant status has any effect on menstrual physiology or symptoms. Dietary intake of antioxidants could help to cope with this oxidative stress and therefore avoid a negative impact on reproductive physiology. A recent systematic review found that increased consumption of fruits and vegetables, fish, and dairy, as sources of many vitamins and minerals (including antioxidants), was associated with less menstrual pain[34].

Unreliable road communications, extreme climate, and cultural heritage all influence food availability in the Andean plateau. Thus, if there truly are differences in menstrual or reproductive health due to hypoxia, these may be overturned by dietary habits and/or traditional medicine alternatives. In fact, in our cohort, nearly 2 out of 3 women who required treatment for menstrual pain preferred traditional herbal remedies over pharmacologic options.

In rural areas, women often face unique challenges due to limited access to medical facilities, education, and resources. Our results highlight some challenges that need to be addressed. Two-thirds of surveyed women did not use contraception 7%), which is similar to published data from 2004 in Peru, where the rate of contraception use was 41%[35]. Barriers to contraception access have been described in several studies and include social, cultural, political, economic, and religious factors[36,37]. These barriers hinder access to sexual and reproductive healthcare, which can worsen women’s health. We observed an unusually low self-reported prevalence of gynecological diseases, but evidence shows that uterine pathology is quite common worldwide[18–20]. This could be due to a lack of assistance from local healthcare services, or to mistrust or low confidence in modern allopathic Western medicine. In our sample, although nearly 1 in 4 women had moderate-severe dysmenorrhea, 77.2% had never sought medical assistance. In addition, when women required analgesia for menstrual pain, 62.3% preferred herbal preparations over modern pharmacological options, and more than 3/4 were self-medicated. This also underscores the importance of indigenous beliefs and the high value of traditional and alternative medicine.

Less than half of the surveyed women regularly visited the obstetrics and gynecology specialist (47.8%), but this may not necessarily reflect a lack of access to women’s healthcare because many preventive actions are carried out by other medical professionals or by non-medical personnel (e.g., cervical cancer screening, contraception).

### Strengths and limitations

The main strength of our study is that it is the first descriptive study to report on the menstrual characteristics of high-altitude residents in the Andes mountain range.

Although we had a rejection rate of 19%, all 32 participants who rejected participation did so in the preliminary stage, before reading any questions; it is unlikely that cultural barriers regarding menstruation were a source of non-response bias.

The study protocol planned to gather data from all Andean countries, but we only obtained data from 3 countries: Chilean women were unavailable, and the COVID-19 pandemic impeded the continuation of the study in Ecuador and Colombia. Thus, our cohort may not be representative of all Andean residents.

The opportunistic sampling method could be a source of bias. Still, we tried to reduce this risk by systematically visiting the same public places in all areas, on weekdays and in the afternoon.

The provision of quality and equal healthcare in South America is complicated. It is a culturally and ethnically diverse region marked by growing economies, high violence rates, political instability, and significant inequality[38]. The healthcare systems here are fragmented, and the geographical adversities of the high Andes aggravate the already challenging delivery of quality care and fairness across society. In this scenario, women are especially vulnerable, and sexual and reproductive healthcare fares worse. Our results highlight the need to improve three key areas: research on the menstrual health of Andean women, access to healthcare in rural high-altitude communities, and the implementation of menstrual education programs. Some initiatives to involve the community in the decision-making process are promising, in Bolivia, for example, but there is a long road ahead to engage everyone and advance sexual and reproductive healthcare for Andean women[39].

Rural women’s health is a vital aspect of healthcare. It addresses unique challenges such as limited access to healthcare facilities, health education, and resources in rural areas. Addressing these issues involves promoting awareness, providing accessible healthcare services, and empowering women with knowledge about their health. Communities, healthcare providers, and policymakers need to collaborate to ensure that women in rural areas receive the care and support they need.

## CONCLUSIONS

Women who live at high altitudes in the Andes (above 3500 m AMSL) have delayed menarche, regular menstrual cycles, and low average menstrual pain (median dysmenorrhea score 3.9/10). However, the prevalence of moderate to severe dysmenorrhea and heavy menstrual bleeding is considerably high and should be studied in more depth. High altitude appears to influence menstrual pain and bleeding; this should be evaluated further in a larger controlled trial comparing sea-level with high Andean residents.

More studies are needed to understand the needs of high-altitude residents in terms of menstrual education and sexual and reproductive healthcare.

## Data Availability

All data produced are available online at figshare

https://doi.org/10.6084/m9.figshare.29876843.v1

## Acknowledgments

We would like to especially thank Betty Choque, MD (Caja Nacional de Salud Potosí, Bolivia), for proofreading and reviewing the survey, and for her collaboration in data acquisition. We also thank Alejandra Sanchez, MD (Clínica del Country, Bogotá, Colombia), for reviewing the survey and study protocol for IRB approval.

## SUPPLEMENTARY MATERIAL

**Table S1: List of study variables**

[+ type of variable]

**Demographics:**

● Age: years [cont.]
● Residence: country [cat.], population [cat.] and altitude [cont.]
● Level of education completed [ord.]: none, elementary, high school [technical or high school], college [university]

**Medical history:**

● Pathological history [dicot.], [cat.]
● Pharmacological history [dicot.], [cat.]
● Smoking [dicot.], [cat.]
● Abdominal surgeries (including cesarean sections) [dicot.], [cat.]
● Physical activity (including frequency) [dicot.], [ord.] and main mode of transportation [cat.]
● Body mass index (BMI) [cont.]

**Gynecologic history:**

● Menarche [cont.]
● Gestations [cont.]
● Abortions [cont.]
● Parity (number of births or cesarean sections) [cont.]
● Contraceptive method [dicot.], [cat.]
● Breastfeeding [dicot.]
● Gynecological check-ups [dicot.]

*Menstruation characteristics:*

● Length of menstrual cycle (days) [cont.]
● Duration of bleeding (days) [cont.]
● Amount of menstrual bleeding: “normal” or possibly heavy (≥1 of 3 statements) – based on the menstrual bleeding questionnaire (MBQ) [ord.]
● Classification of dysmenorrhea into grades (0-3). [ord.]
● Level of dysmenorrhea according to graphic rating scale GRS (score 0 to 10) [cont.]
● Previous consultation for dysmenorrhea [dicot.], [cat.]
● Analgesia for dysmenorrhea [dicot.], [cat.] + type of analgesia [cat.]

**Figure S2: Study survey (orginal in Spanish)**

See supplementary PDF file. The original survey was written in Spanish; an English translation can be found here: https://bit.ly/HIRU-high-altitude-menstruation-survey

**Table S3: STROBE checklist**

STROBE Statement—checklist of items that should be included in reports of observational studies

See supplementary PDF file.

**Figure S4:**
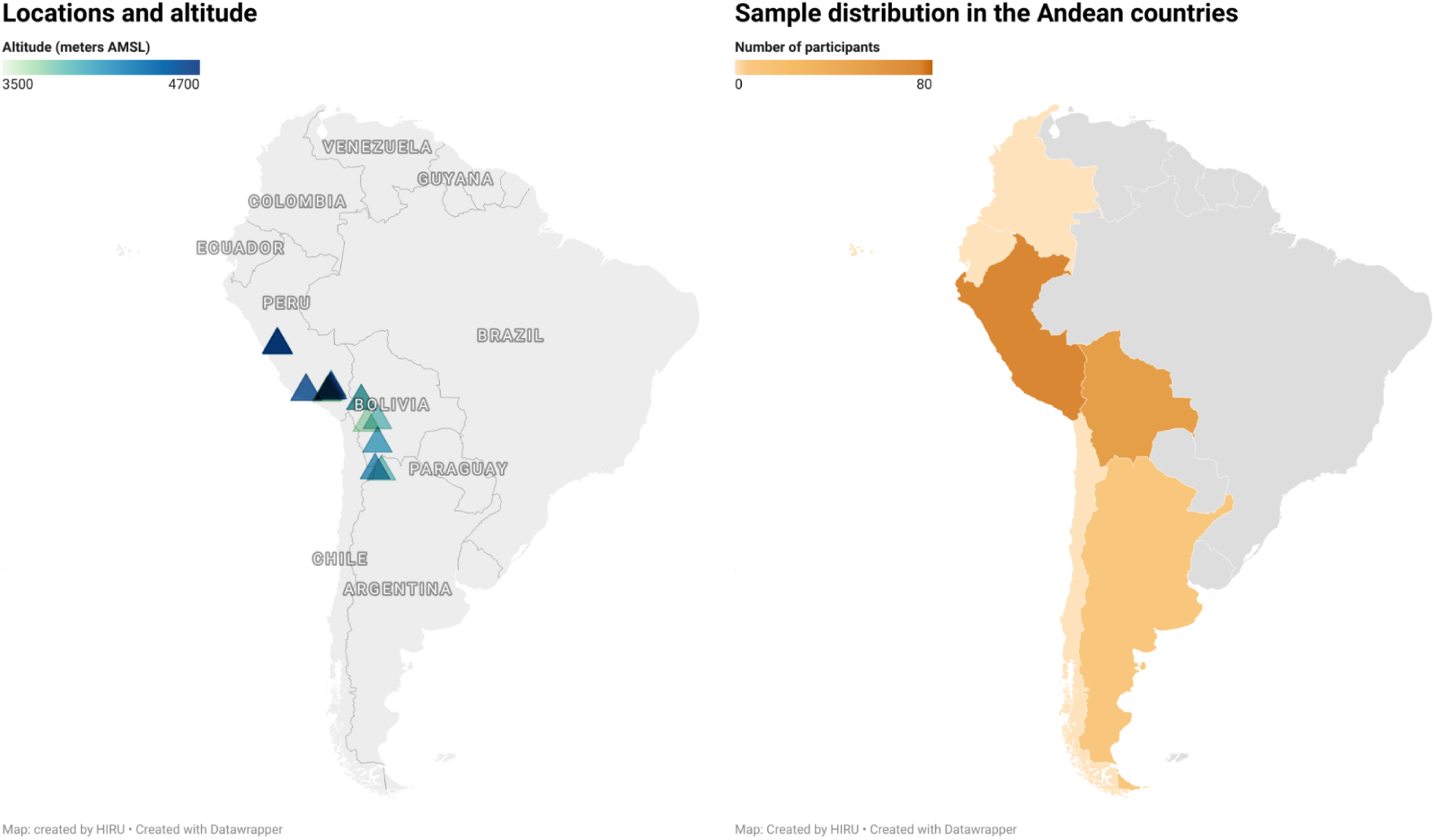
Locations, altitude, and distribution of the sample. Interactive maps can be accessed online: Locations and altitude: https://www.datawrapper.de/_/MgvBR/?v=2 Sample distribution: https://www.datawrapper.de/_/Fff8a/?v=2

