## Supplementary material S2 for "Menstruation at high altitude: results from an opportunistic cross-sectional survey of Andean women living above 3500 m"

### INFORMATION FOR PARTICIPANTS

Research project entitled: "*Study of dysmenorrhea in women living at high altitude*".

Principal Investigators: Alejandro Correa Paris, Verónica Gorraiz Ochoa
Organization: HIRU - Health Independent Research United

Objectives:
We request your participation in this research project whose main objective is to deepen the knowledge of pain during menstruation or menstrual period (called dysmenorrhea) in women living at high altitude (equal or higher than 3500 m above sea level).

Benefits:
There may be no direct benefit from your participation in this study. However, a deeper understanding of the symptoms may help in the identification of diseases. This could in the future benefit other women suffering from symptoms or related diseases, and thus contribute to a better understanding and treatment of these diseases.

Procedure:
The study will be conducted by means of an anonymous survey that will provide data to be analyzed and subsequently published in the medical literature.

Risks:
Your participation is voluntary and carries no risk.

Protection of personal data:
The personal data collected will be those necessary for the purposes of the study. Any information of a personal nature that may be identifiable will be kept by computerized methods under secure conditions by those responsible for the study. Access to such information will be restricted to designated HIRU personnel or other authorized personnel who will be obliged to maintain the confidentiality of the information.

In accordance with current legislation, you have the right to be informed of the results of the study. If you wish to know this information, you may contact the organization by e-mail.

Your participation in the study is completely voluntary, and you may decline at any time.

### A study of dysmenorrhea in women living at high altitude

**ELIGIBILITY CRITERIA:**
- ☐ I am between 16 and 45 years old.
- ☐ I am not currently pregnant.
- ☐ I usually have menstruation.
- ☐ I have received information and agree to participate in the survey.

Please answer the following questions.

Please take the time to read each question.

Some questions require you to fill in the blanks ( ____ ) by typing in your answer. Others require you to put a ✗ in the box that best describes your answer.

If you do not understand a question, you may ask the interviewer for clarification.

***Thank you for taking the survey!***

#### Sociodemographic data

1. Date of birth (dd/mm/yyyy):
   ___ / ___ / _______

2. Completed level of studies:
   - ▢ None
   - ▢ Primary
   - ▢ Secondary school (technical or bachelor's degree)
   - ▢ Higher (university)

3. Residence (City, Country):
   _______________ , _____________

#### Medical history

4. Do you have or do you suffer from any disease?
   ▢ No ▢ Yes. Specify which one(s): _____________

5. Do you regularly take any type of medication, remedy or medicine?
   ▢ No ▢ Yes. Specify:
   - Prescribed by physician. Specify: _____________
   - Self-medicated (herbs or home remedies). Specify: ___________

6. Do you smoke?
   ▢ No ▢ Yes

7. Have you ever had an operation or surgery on your abdomen or belly?
   ▢ No ▢ Yes. Specify which one(s): _________________________

8. Do you engage in any type of physical activity or sport?
   ▢ No ▢ Yes. How many days per week (on average)? _____

9. How do you usually get around to perform your daily activities?
   ▢ Walking ▢ Bicycle ▢ Vehicle (own/public transportation)

10. Indicate your weight and height:
    Weight (in kg or pounds): _____ Height (in meters): _____ ▢ Don't know

#### Gynecological history

11. How old were you when you first got your period? _____

12. How many times have you been pregnant (including miscarriages or losses)? _____

13. Have you ever had a pregnancy loss or miscarriage?
    ▢ No ▢ Yes. Specify how many: _____

14. How many children have you had in total (births or cesarean sections)? _____

15. Do you use any method to prevent pregnancy (contraceptive method)?
    ▢ No ▢ Yes. Specify which: ______________

16. Are you currently breastfeeding or nursing?
    ▢ No ▢ Yes

17. Do you periodically go to the gynecologist for a routine check-up?

▢ No ▢ Yes

**Characteristics of menstruation**

18. Approximately how often do you get your period?
    ▢ 21 days or less    Between 21 and 35 days    ▢ More than 35 days

19. How many days does your menstrual bleeding last?
    ▢ 7 days or less    ▢ Between 8 and 14 days    ▢ More than 14 days

20. Regarding the **amount of bleeding** during menstruation, check the boxes that describe your case (one or more):
    ▢ The amount of bleeding is normal and does not affect daily activities.
    ▢ I need to change protection (towel, pad or tampon) every hour or less.
    I need to use more than one protection (double towel, diaper, etc.).
    ▢ I need to get up at night to change protection because of bleeding.

21. Regarding any **pain** you may have during menstruation, check the box that most closely matches your case (only one):

| | ***Features*** | ***Work/school ability*** | ***Other symptoms*** | ***Pain medication or remedies*** |
|---|---|---|---|---|
| ▢ | It is not painful and does not affect daily activities. | Not affected | No | Does not require |
| ▢ | Painful but does not impede normal activities. Mild pain. | Rarely affected | No | Occasionally |
| ▢ | Pain is moderate and affects daily activities. You can work or go to school if you take medications or remedies. | Moderately affected | Few | Always |
| ▢ | The pain is intense and prevents daily activities. | Very much affected | Evident (nausea, vomiting, fatigue, etc.) | Not very effective |

22. On the line below, draw a line through the point that corresponds to the **pain** you usually experience with your period or menstrual period:

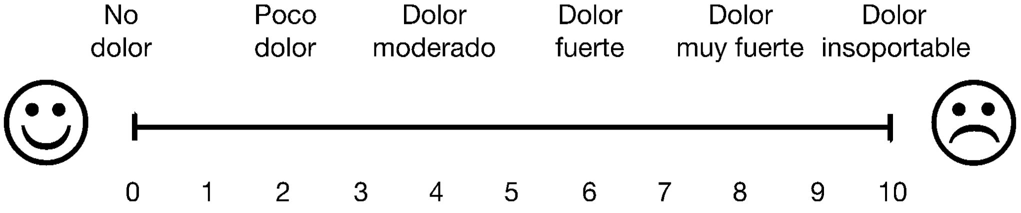


23. Have you ever consulted a physician for menstrual pain?
    ▢ No ▢ Yes

24. Do you usually take any type of medication, remedy or medication for menstrual pain?

▢ No ▢ Yes. Specify:

- Prescribed by physician. Specify: __________
- Self-medicated (herbs or home remedies). Specify: __________
