## Supplementary material S3 for "Menstruation at high altitude: results from an opportunistic cross-sectional survey of Andean women living above 3500 m"

STROBE Statement—checklist of items that should be included in reports of observational studies

| | Item No. | Recommendation | Page No. | Relevant text from manuscript |
|---|---|---|---|---|
| **Title and abstract** | 1 | (*a*) Indicate the study's design with a commonly used term in the title or the abstract | 1 | *opportunistic cross-sectional survey* |
| | | (*b*) Provide in the abstract an informative and balanced summary of what was done and what was found | 2-3 | See full abstract |
| **Introduction** | | | | |
| Background/rationale | 2 | Explain the scientific background and rationale for the investigation being reported | 4 | See introduction |
| Objectives | 3 | State specific objectives, including any prespecified hypotheses | 5 | *to describe the characteristics of the menstrual cycle and the prevalence of dysmenorrhea in residents living above 3500 m (...) in the Andean mountains.* |
| **Methods** | | | | |
| Study design | 4 | Present key elements of study design early in the paper | 5 | *We conducted an opportunistic cross-sectional survey* |
| Setting | 5 | Describe the setting, locations, and relevant dates, including periods of recruitment, exposure, follow-up, and data collection | 5 | *between August 2019 and March 2020 in all available and permanently inhabited settlements above 3500 m above mean sea level (AMSL) in the* |

| | | | | |
|---|---|---|---|---|
| | | | | *South American Andes mountain range* |
| Participants | 6 | (*a*) *Cohort study*—Give the eligibility criteria, and the sources and methods of selection of participants. Describe methods of follow-up<br><br>*Case-control study*—Give the eligibility criteria, and the sources and methods of case ascertainment and control selection. Give the rationale for the choice of cases and controls<br><br>*Cross-sectional study*—Give the eligibility criteria, and the sources and methods of selection of participants | 6 | *We included all women of reproductive age who were permanent residents at an altitude greater than or equal to 3500 m AMSL. Permanent residents were defined as having lived in that location for at least 6 months (...) The inclusion criteria were: age between 16 and 45 years, and having a menstrual period. The exclusion criteria were: amenorrhea (>3 months), current pregnancy, menopause, and inability to complete the survey.* |
| | | (*b*) *Cohort study*—For matched studies, give matching criteria and number of exposed and unexposed<br><br>*Case-control study*—For matched studies, give matching criteria and the number of controls per case | NA | NA |
| Variables | 7 | Clearly define all outcomes, exposures, predictors, potential confounders, and effect modifiers. Give diagnostic criteria, if applicable | 7 | *main outcome measure was the dysmenorrhea intensity score. (…) the severity of dysmenorrhea (grades 0 to 3, see Figure 4) based on the patient’s symptoms. The secondary outcomes included: other characteristics of dysmenorrhea (i.e., need for* |

| | | | | |
|---|---|---|---|---|
| | | | | *analgesia), menstrual bleeding and cycle characteristics, lifestyle and anthropometric characteristics, including the frequency of physical activity and main mode of transportation, past medical and gynecological history, and demographic characteristics, including age and education* |
| Data sources/ measurement | 8* | For each variable of interest, give sources of data and details of methods of assessment (measurement). Describe comparability of assessment methods if there is more than one group | 7 | *We used the validated 10-grade graphic rating scale (GRS) to determine the dysmenorrhea intensity score (0 to 10), (...) We also used a verbal multidimensional system to classify the severity of dysmenorrhea (grades 0 to 3, see Figure 4) based on the patient's symptoms.*<br>See supplementary material: Table S1 |
| Bias | 9 | Describe any efforts to address potential sources of bias | 7 | *We used the validated 10-grade graphic rating scale (GRS) to determine the dysmenorrhea intensity score (0 to 10), which is easier to understand than the traditional visual analog scale (VAS) and yields fewer* |

| | | | | |
|---|---|---|---|---|
| | | | | *erroneous interpretations and false 0 values* |
| Study size | 10 | Explain how the study size was arrived at | 8 | *The sample size was calculated in advance using the Granmo calculator software (v7.12) and based on the results of a study describing the prevalence and severity of dysmenorrhea[16] where the mean score was 6.79 ± 2.64 on a 10-grade VAS. On the basis of these results, we determined that a random sample of 119 individuals would be sufficient to estimate the population mean with a precision of ± 0.5, standard deviation of 2.64, and 95% confidence* |

Continued on next page

| | | | | |
|---|---|---|---|---|
| Quantitative variables | 11 | Explain how quantitative variables were handled in the analyses. If applicable, describe which groupings were chosen and why | 8 | *For the descriptive analysis, we calculated percentages, medians, interquartile range (IQR), and standard error of the mean (SEM) when appropriate.* |
| Statistical methods | 12 | (*a*) Describe all statistical methods, including those used to control for confounding | 8 | *We used statistical tests as per the distribution of data (normal vs. non-normally distributed), the Chi-square or Fisher tests for qualitative variables, and the Mann-Whitney U or t-tests based on the nature of the quantitative variables.* |
| | | (*b*) Describe any methods used to examine subgroups and interactions | NA | NA |
| | | (*c*) Explain how missing data were addressed | 7 | *Incomplete surveys (if more than 80% of questions were left unanswered) were excluded from the analysis* |
| | | (*d*) *Cohort study*—If applicable, explain how loss to follow-up was addressed<br>*Case-control study*—If applicable, explain how matching of cases and controls was addressed<br>*Cross-sectional study*—If applicable, describe analytical methods taking account of sampling strategy | NA | NA |
| | | (*e*) Describe any sensitivity analyses | NA | NA |
| **Results** | | | | |

| | | | | |
|---|---|---|---|---|
| Participants | 13* | (a) Report numbers of individuals at each stage of study—eg numbers potentially eligible, examined for eligibility, confirmed eligible, included in the study, completing follow-up, and analysed | 9 | See figure 1.<br>*The response rate was 81% (136/168); most answers were collected from written surveys (95%), and we did not exclude any surveys because of missing data. (...) We analyzed a total of 136 participants.* |
| | | (b) Give reasons for non-participation at each stage | | See figure 1 |
| | | (c) Consider use of a flow diagram | | See figure 1 |
| Descriptive data | 14* | (a) Give characteristics of study participants (eg demographic, clinical, social) and information on exposures and potential confounders | 9-12 | See Results section & Tables 1 and 2. |
| | | (b) Indicate number of participants with missing data for each variable of interest | | See figure 1 |
| | | (c) *Cohort study*—Summarise follow-up time (eg, average and total amount) | NA | NA |
| Outcome data | 15* | *Cohort study*—Report numbers of outcome events or summary measures over time | NA | NA |
| | | *Case-control study*—Report numbers in each exposure category, or summary measures of exposure | NA | NA |
| | | *Cross-sectional study*—Report numbers of outcome events or summary measures | *9-12* | See Results section & Tables 1 and 2. |
| Main results | 16 | (*a*) Give unadjusted estimates and, if applicable, confounder-adjusted estimates and their precision (eg, 95% confidence interval). Make clear which confounders were adjusted for and why they were included | 9-12 | *The median dysmenorrhea score was 3.9 (IQR 2.8), with an estimated population mean of 3.7 ± 0.4.*<br>See Results section |
| | | (*b*) Report category boundaries when continuous variables were categorized | NA | NA |

| | | | |
|---|---|---|---|
| | (*c*) If relevant, consider translating estimates of relative risk into absolute risk for a meaningful time period | NA | NA |

Continued on next page

| | | | | |
|---|---|---|---|---|
| Other analyses | 17 | Report other analyses done—eg analyses of subgroups and interactions, and sensitivity analyses | NA | NA |
| **Discussion** | | | | |
| Key results | 18 | Summarise key results with reference to study objectives | 12 | *We found that women of reproductive age residing at high altitudes in the Andes have mostly regular menstrual cycles. The average menstrual pain score was medium-low (median 3.9, IQR 2.8), but 23.6% of participants had moderate to severe dysmenorrhea* |
| Limitations | 19 | Discuss limitations of the study, taking into account sources of potential bias or imprecision. Discuss both direction and magnitude of any potential bias | 16 | *The opportunistic sampling method could be a source of bias* |
| Interpretation | 20 | Give a cautious overall interpretation of results considering objectives, limitations, multiplicity of analyses, results from similar studies, and other relevant evidence | 17 | *The healthcare systems here are fragmented, and the geographical adversities of the high Andes aggravate the already challenging delivery of quality care and fairness across society. In this scenario, women are especially vulnerable, and sexual and reproductive healthcare fares worse.* |
| Generalisability | 21 | Discuss the generalisability (external validity) of the study results | 17 | *Our results highlight the need to improve three main areas: research on Andean women's menstrual health, access to healthcare in rural high-altitude communities, and implementation of menstrual education programs.* |

| | | | | |
|---|---|---|---|---|
| **Other information** | | | | |
| Funding | 22 | Give the source of funding and the role of the funders for the present study and, if applicable, for the original study on which the present article is based | 1 | *The present study did not receive any grants or funding.* |

*Give information separately for cases and controls in case-control studies and, if applicable, for exposed and unexposed groups in cohort and cross-sectional studies.

**Note:** An Explanation and Elaboration article discusses each checklist item and gives methodological background and published examples of transparent reporting. The STROBE checklist is best used in conjunction with this article (freely available on the Web sites of PLoS Medicine at http://www.plosmedicine.org/, Annals of Internal Medicine at http://www.annals.org/, and Epidemiology at http://www.epidem.com/). Information on the STROBE Initiative is available at www.strobe-statement.org.
